# Linkage analysis identifies an isolated strabismus locus at 14q12 overlapping with FOXG1 syndrome region

**DOI:** 10.1101/2020.04.24.20077586

**Authors:** Xin (Cynthia) Ye, Nicole M. Roslin, Andrew D. Paterson, Care4Rare Canada, Christopher Lyons, Victor Pegado, Phillip Richmond, Casper Shyr, Oriol Fornes, Xiaohua Han, Michelle Higginson, Colin J. Ross, Deborah Giaschi, Cheryl Y. Gregory-Evans, Millan Patel, Wyeth W. Wasserman

## Abstract

Strabismus is a common condition, affecting 1-4% of individuals. Isolated strabismus has been studied in families with Mendelian inheritance patterns. Despite the identification of multiple loci via linkage analyses, no specific genes have been identified from these studies. The current study is based on a seven-generation family with isolated strabismus inherited in an autosomal dominant manner. A total of 13 individuals from a common ancestor have been included for linkage analysis, and a single linkage signal has been identified at chromosome 14q12 with a multipoint LOD score of 4.69. Disruption of this locus is known to cause FOXG1 syndrome (or congenital Rett syndrome; OMIM #613454 and *164874), in which 84% of affected individuals present with strabismus. With the incorporation of next generation sequencing and in-depth bioinformatic analyses, a 4bp non-coding deletion was prioritized as the top candidate for the observed strabismus phenotype. The deletion is predicted to disrupt regulation of *FOXG1*, which encodes a transcription factor of the Forkhead family. Suggestive of an auto-regulation effect, the disrupted sequence matches the consensus FOXG1 and Forkhead family transcription factor binding site and has been observed in previous ChIP-seq studies to be bound by Foxg1 in early mouse brain development. The findings of this study indicate that the strabismus phenotype commonly observed within FOXG1 syndrome is separable from the more severe syndromic characteristics. Future study of this specific deletion may shed light on the regulation of *FOXG1* expression and may enhance our understanding of the mechanisms contributing to strabismus and FOXG1 syndrome.

**Author summary:** Eye misalignment, or strabismus, can affect up to 4% of individuals. When strabismus is detected early, intervention in young children based on eye patching and/or corrective lenses can be beneficial. In some cases, corrective surgeries are used to align the eyes, with many individuals requiring multiple surgeries over a lifetime. A better understanding of the causes of strabismus may lead to earlier detection as well as improved treatment options. Hippocrates observed that strabismus runs in families over 2,400 years ago, an early recognition of what we now recognize as a portion of cases arising from genetic causes. We describe a large family affected by strabismus and identify a single region on chromosome 14 that may be responsible. The region contains *FOXG1*, in which mutations are known to cause a severe syndrome, with 84% of affected individuals also having strabismus. We identify a 4bp deletion in the region that appears to auto-regulate when *FOXG1* is active. Future study of this genetic alteration may enhance our understanding of the mechanisms of strabismus.

## Introduction

Strabismus, also known as crossed eyes or squint, affects 1-4% of individuals. Diagnosis and treatments for strabismus are well-established, but the pathophysiology for most isolated strabismus remains largely unknown. Disturbances anywhere along the visual sensory or the oculomotor pathways can be postulated to lead to eye deviation (1).

As early as Hippocrates’ time, strabismus was recognized as a genetic disorder based on an observation of its tendency to cluster within families (2). During the last century, twin and family studies have demonstrated a substantial genetic contribution to strabismus, and both autosomal dominant and autosomal recessive transmission patterns have been reported (3). Recently a genome wide association study reported two variants with small effect sizes (4). Strabismus occurs commonly as one phenotype amongst many in syndromes, such as congenital Rett syndrome (FOXG1 syndrome) and Joubert syndrome, in which 84% and 75% of individuals display a strabismus phenotype respectively (5,6). On the other hand, families displaying isolated strabismus transmitting in simple Mendelian patterns are uncommon. In a few such families, genetic loci on chromosomes 4, 6, 7, 12, 16, 19 have been identified for isolated strabismus, but no causal gene has been identified in these regions (3). Not overlapping the reported loci, eleven genes (*PHOX2A, ROBO3, KIF21A, <u>SALL1</u>, TUBB3, <u>HOXB1</u>, <u>SALL4</u>, CHN1, <u>HOXA1</u>, TUBB2B, <u>MAFB</u>*) (7), of which five encode transcription factors (<u>underlined</u>), have been identified for a subgroup of strabismus associated with congenital cranial dysinnervation disorders (7), but the genetic etiology of other strabismus subtypes remains elusive. Identification of a locus with high confidence, determination of a causal gene and detailed mechanistic insights at the nucleotide level would provide new insights into the molecular mechanisms of strabismus.

We compiled a large, seven-generation, non-consanguineous pedigree with 21 individuals exhibiting isolated strabismus, consistent with an autosomal dominant inheritance pattern. Through genome-wide linkage analysis of 8 affected and 4 unaffected individuals from one branch and one affected individual from a separate branch, we mapped this familial strabismus to chromosome 14q12, which overlaps with the FOXG1 syndrome locus. Next generation sequencing and in-depth analysis identified a strong candidate deletion variant within this locus. The data supports that the deletion localizes to a FOXG1 transcription factor binding site (TFBS), suggesting an auto-regulatory loop may be disrupted that controls *FOXG1* expression in early brain development.

## Results

### Pedigree and participant profile

A seven-generation pedigree of European origin, with over 176 individuals, including those who were deceased, was compiled. Individuals were labelled as strabismic either through medical records or strong family anecdotes. A roughly even distribution of strabismus cases was observed between females (12 individuals) and males (nine individuals) and the disorder was transmitted both maternally and paternally. Among the three extensively traced branches (Figure 1a), branch 1 was most well-documented and contained most of the participants in this study. In branch 1, strabismus was reported across four consecutive generations. An autosomal dominant inheritance model with high penetrance best matched the qualitative observation.

**Figure 1.**
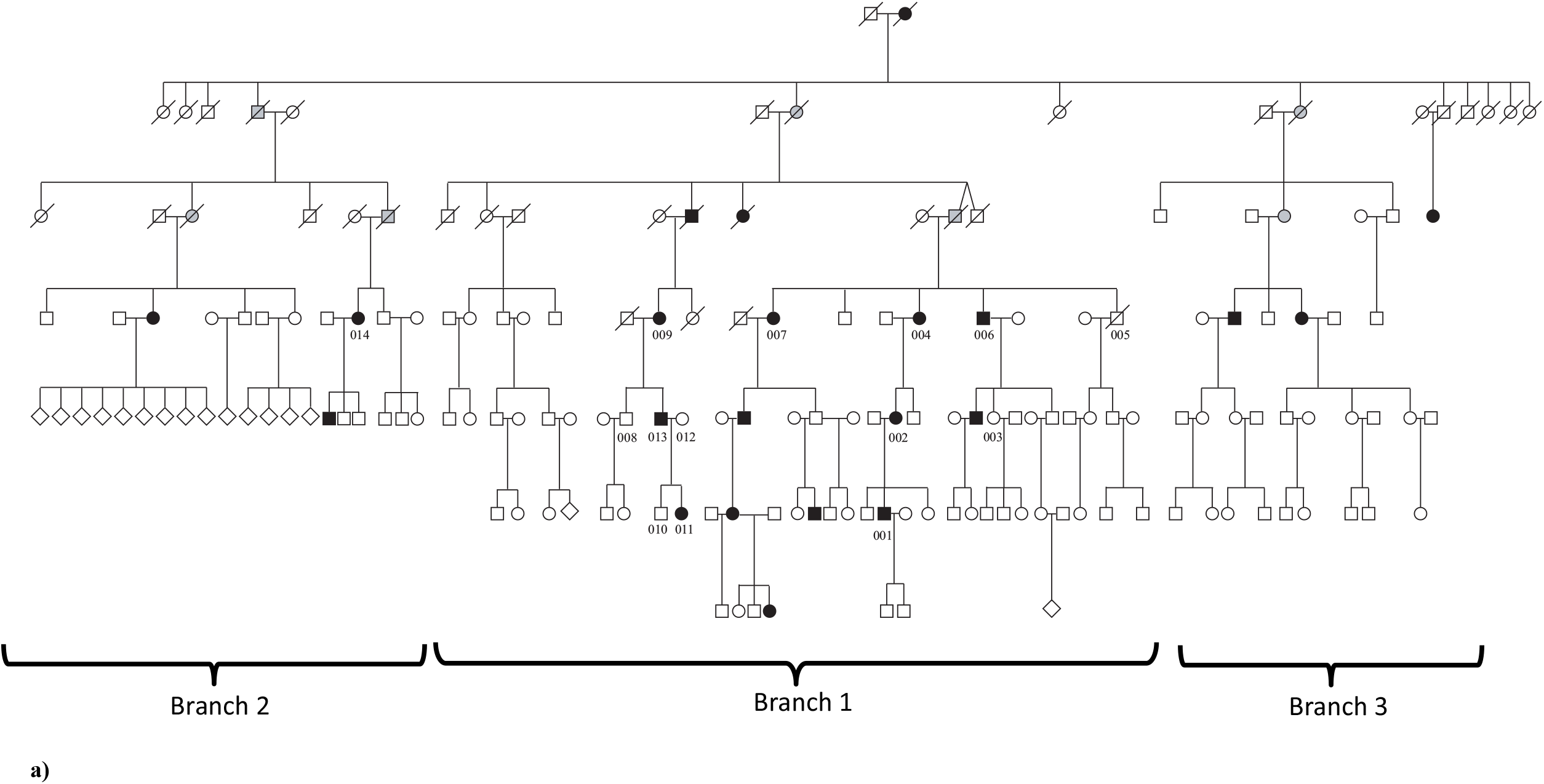
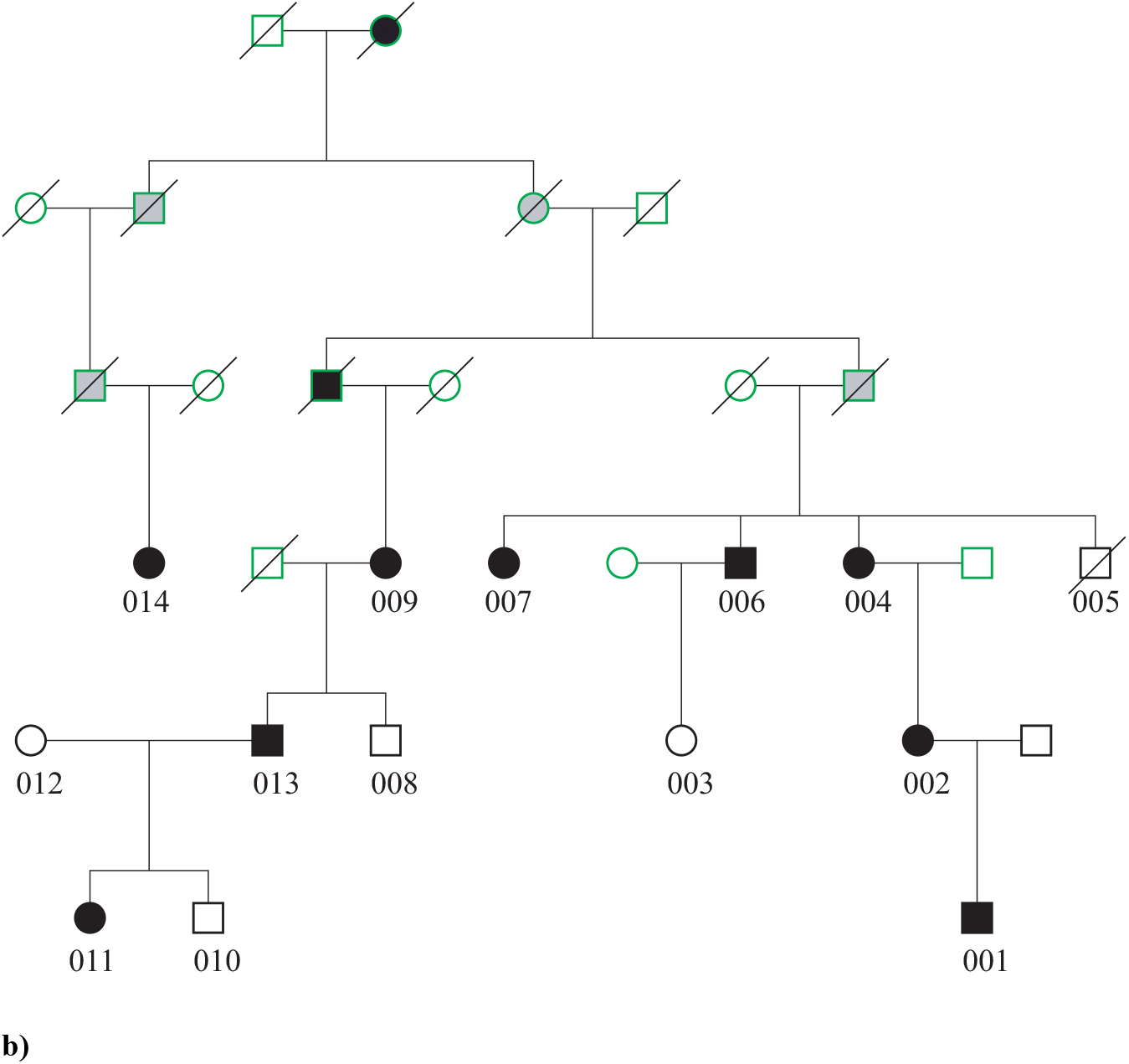
Pedigree for the subject family with isolated strabismus. a) The pedigree represents a seven-generation family with 176 individuals, including deceased individuals. Three major branches are identified: 12 study participants come from Branch 1 and one (014) comes from Branch 2. Individual 012 was not included in the linkage analysis. Black represents affected individuals, white represented unaffected individuals, and grey represents obligate carriers. b) **Simplified Branch 1 of the subject family showing the genotyped individuals (with study ID) and ancestors required to link them. Individual 014 represents Branch 2, and all the other individual comes from Branch 1**. Individuals whose status was not confirmed clinically were coded as unknown for the linkage analysis. These individuals were indicated by green outlines.

To characterize the strabismus in this family, each of the nine affected descendants indicated in Figure 1b was seen by one of three participating ophthalmologists specialized in strabismus. All individuals underwent complete ophthalmic examination with attention to ocular motility both before and after pharmacologic cycloplegia. The specific characteristics of strabismus were not uniform across the descendants in the family (Table 1). The strabismus phenotypes could be grouped into two broad directional categories: esotropia (an eye turns in) and hypertropia (an eye turns up). Both esotropia and hypertropia were noted in individual 014, but this individual had undergone multiple corrective surgeries and childhood medical records were not available. On the other hand, individuals 011, 013 and 009 are in consecutive generations and had no history of extraocular muscle surgery. Individual 011 presented with esotropia while the other two displayed hypertropia (009, 013). The unaffected status was confirmed by past medical history provided by the participating family members. In the clinical ophthalmology exams and in oral reports from subjects, there were no other phenotypes observed broadly in the individuals with strabismus.

**Table 1.** Specific characteristics of strabismus across the affected descendants in the subject family.

| Identifier | Type of reported strabismus | Age of onset | Severity | Eye movement full? | Concomitant? | Stereopsis | Strabismus surgery? | Intraocular pressure | Optic disc and macula | Other ocular conditions |
| --- | --- | --- | --- | --- | --- | --- | --- | --- | --- | --- |
| 001 | Accommodative left esotropia with an A pattern | 9 months | Severe | Y | Incomitant | Absent | bilateral strabismus surgery at age two and at age three; botulinum toxin injection at the age of 4 | 14 mm Hg OU | Normal | Mild amblyopia, left eye |
| 002 | Hypotropia and left exotropia. Consecutive exodeviation after initial esotropia surgery. | unclear | Severe | Y | NA | Absent | strabismus surgery at ages 3,8, and 16 | 16 mm Hg OU | Normal | No diplopia, suppression |
| 004 | Right esotropia | age of 3 | Severe | Y | Concomitant | NA | strabismus surgery at ages 7 and 10 | 16 mm Hg OU | Normal | right cataract, right dense amblyopia |
| 006 | NA | NA | NA | NA | NA | NA | NA | NA | NA | NA |
| 007 | Hypertropia | unclear | cannot be ascertained | Y | cannot be ascertained | cannot be ascertained | NA | 18 mm Hg OU | Optic disc normal; macular degeneration on the left eye and macular drusen on the right eye | cataract, diplopia, latent nystagmus, left eye suppression |
| 009 | Congenital right hypertropia; excyclotorsion | after 2, exact onset time unclear, aware of ocular misalignment at the age of 9 | Mild | Y | Incomitant | Intact | N | 16 mm Hg OU | Normal | Presbyopia |
| 011 | Childhood left esotropia | before the age of 2 | Moderate | Y | Concomitant | Gross fusion & stereopsis | N | NA | Normal | Myopia |
| 013 | Congenital right hypertropia | unclear, aware of ocular misalignment at the age of 6 | Mild | Y | Incomitant | Have the ability to use the 2 eyes together with stereopsis potential when the 2 images are artificially aligned | N | NA | Myopic optic discs | Myopia |
| 014 | Esotropia, right hypertropia; excyclotorsion in both eyes | at birth | Severe | Y | Incomitant | Absent | 2 strabismus surgeries during 50s | NA | Normal | cataract, diplopia, latent nystagmus, left eye suppression |

### Linkage analysis and haplotype analysis

Initial linkage analysis was performed with 12 family members; 8 affected and 4 unaffected individuals (excluding 012 and 014 from Figure 1b). Simulations under the alternative hypothesis (linkage) generated a maximum LOD score of 3.56, under an autosomal dominant model with disease allele frequency q = 0.005, 99% penetrance, and 0.2% phenocopy rate. The LOD score curves did not change significantly across a range of disease allele frequency settings (results not shown).

With the same linkage analysis parameters as in the simulation, the largest LOD score based on the observed genotypes was 3.55, close to the simulated maximum. This chromosome 14 locus was the only region with a LOD score higher than 3, and exceeds the standard threshold for genome-wide significance of 3.3 (8). This linked region spanned approximately 10 Mb and was bounded by rs7146411 and rs1951187, corresponding to chr14: 22,779,843 - 32,908,192 (hg19). Chromosome 14q12 is therefore a novel locus for isolated strabismus.

Individual 014 representing branch 2 in the pedigree was recruited to the study after the genome-wide genotyping was performed. An expanded linkage analysis with corresponding SNPs extracted from 014’s whole genome sequencing (WGS) data further supported the linkage to chromosome 14. Simulations under the alternative hypothesis (linkage) generated a maximum LOD score of 4.69 for the same region, under an autosomal dominant model with disease allele frequency q = 0.005, 99% penetrance, and 0.2% phenocopy rate. We observed a LOD score of 4.69, and the region remained as the sole candidate (Figure 2a). In addition, we performed non-parametric linkage analyses and identified the same linkage region (Figure 2b).

**Figure 2.**
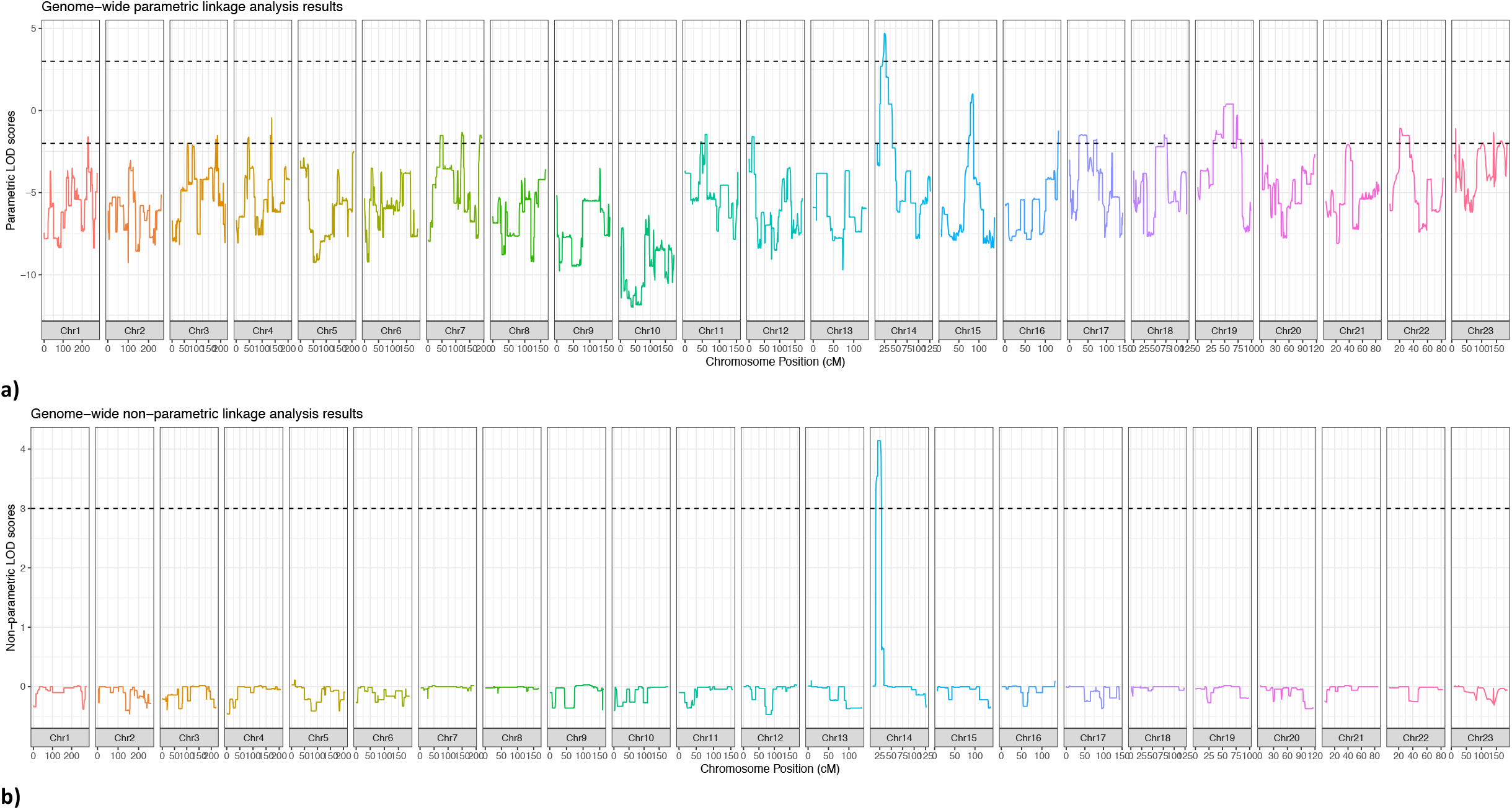
Linkage analysis for subject family. a) Parametric analysis. An expanded linkage analysis was performed in all 13 individuals who shared the common ancestor. We observed a LOD score of 4.69 for the linkage region in chromosome 14. The top dashed line indicates a LOD of 3 and the bottom dashed line indicates a LOD of -2. **b)** Non-parametric analysis. We performed non-parametric analyses and obtained the same linkage region on chromosome 14. The dashed line indicates a LOD of 3

Haplotype analysis was used to complement linkage analysis by providing visual confirmation of the statistical testing. An approximately 8.5 Mb region (chr14:22,779,843 - 31,289,720) was shared between the nine affected descendants of the common ancestor. An unaffected descendant (005, subsequently deceased) shared a 5.5 Mb region within the linked region (Figure 3a and Figure 3b). Thus, an approximately 3 Mb region was shared exclusively by nine affected descendants, corresponding to chr14: 28,467,136 - 31,289,720. The LOD score was above 4.60 for chr14:28,467,136 – 30,045,978 (Supplementary Figure 1). The 3 Mb region situates within a gene poor region (Figure 3b).

**Figure 3.**
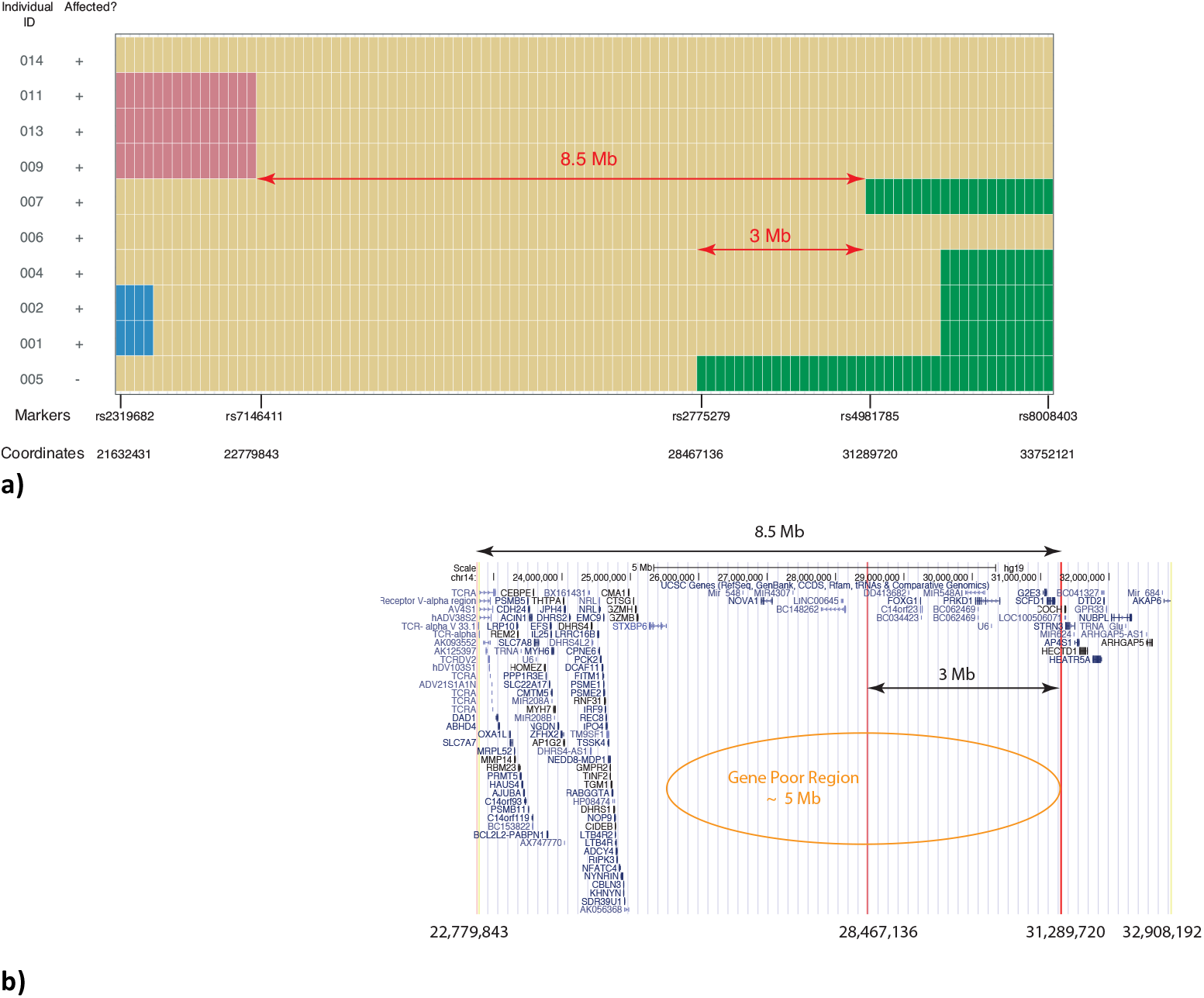
Linkage region. a) Haplotype analysis for subject family. Each row represents an individual, and each column represents a marker used in the linkage analysis. The markers displayed span chr14:22,779,843 – 31,289,720 (∼8.5 Mb) shared between nine affected individuals. In addition to the nine affected, an unaffected individual is included who shares a 5.5 Mb portion of the region. Thus, an approximately 3 Mb region was shared exclusively by 9 affected descendants, corresponding to chr14: 28,467,136 – 31,289,720 (hg19). Yellow indicates the haplotype inherited from the common ancestor. Each of the other colors indicate a different haplotype from a different ancestor. The 8.5 Mb and the core shared ∼3 Mb are indicated. For clarity, unaffected individuals not sharing a portion of the region with those affected are not displayed. b) Genes across the linkage region. The genes reported in the UCSC gene track from the UCSC Genome Browser are displayed for the linkage region. The ∼8.5 Mb region, and the core ∼3 Mb region are indicated. The core ∼3 Mb region lies within a gene poor region.

### No impactful coding variant in the 10 Mb locus identified through WES and WGS

Whole exome sequencing (WES) showed that two affected third-degree cousins (001 and 011) shared 119 heterozygous non-synonymous variants across the entire exome. A subset of 60 variants among the 119 had a frequency lower than 1% in an in-house database of a rare-disease-WES project (9). Only one of the variants (chr14: 31061628 A>G, rs145527124) was located within the 10 Mb locus, falling within exon 5 of *G2E3* (G2/M-phase specific E3 ubiquitin protein ligase) (NM_017769.4).

This variant leads to Ile113Val alteration in ENST00000206595. In gnomAD v 2.1.1 European (non-Finnish) population, there are two homozygotes for this variant and the overall allele frequency is 0.17% and 2 homozygotes while gnomAD v3 has an overall allele frequency of 0.15% and another homozygote (10). In addition, in the 1000 Genomes project (11) the allele frequency in Punjabi from Lahore, Pakistan (PJL) is 1.6%. This *G2E3* variant was not supported as a candidate by computational analysis (predicted to be “tolerated” with SIFT (12) and “benign” with Polyphen (13)). Qualitative review of the literature did not suggest a potential role for *G2E3* in a strabismus phenotype.

The lack of candidate variants from WES motivated the generation of WGS for individuals 001, 013, and 014, who were selected to represent the three branches of the pedigree. The only coding variant detected with a frequency of ≤ 1% in the 10 Mb region was the aforementioned *G2E3* variant.

### WGS and bioinformatics analyses highlight a heterozygous non-coding variant in a regulatory region of *FOXG1*

Our analyses showed that the WGS on 001, 013, and 014 did not capture some low complexity regions, raising concern that the protocol used at the time of generation might fail to detect repetitive sequences and small genomic structural alterations. As current WGS protocols could better detect such properties, we generated WGS for an affected parent-child trio (011, 012, and 013). For simplicity, we report variants from the trio WGS set for comprehensiveness in the following sections.

Within the 3 Mb region, a total of 664 variants were shared by both affected individuals (based on a dominant model of transmission). No copy number variations (CNV) were detected. We focused on variants with a frequency ≤ 1% and which have been reported in fewer than 10 homozygotes in gnomAD 2.0, criteria met by 24 of the 664 variants. The only coding variant of these 24 was the *G2E3* coding variant reported above. (These potentially identifying variants are available from the corresponding author upon request and under an appropriate data handling agreement.)

As 23 of 24 candidate variants prioritized in the WGS analysis were non-coding, we used diverse methods to annotate non-coding variants with regulatory information. There is no standard practice to annotate non-coding variants, so we used a variety of approaches to identify those within potential regulatory elements. One variant was noted recurrently as interesting using a variety of bioinformatic predictions. Among the 23 candidate variants, only chr14:29247628 TAAAC>T (Supplementary Figure 2) has been assigned a CADD score over 20 (14) and is ∼10 kb 3’ from *FOXG1*. This variant is situated within a potential regulatory region, as suggested by the presence of DNase-seq peaks and the histone marks H3K4me1, H3K4me2, H3K4me3, H3K9ac and H3K27ac, which are associated with promoters/enhancers (15). This deletion was absent in gnomAD v 2.1.1 and v 3 and dbSNP build 152 and 153 (16). This deletion was confirmed through Sanger sequencing to be present in all nine affected subjects and none of the unaffected individuals.

Since the upper limit of the reported prevalence of strabismus is 4%, additional variants with frequency >1% and ≤4% in gnomAD were also obtained for examination. A total of 54 additional variants were identified in the candidate region. However, none of these variants situated on protein coding regions or were assigned a CADD score over 20.

We examined topologically associating domains (TADs) for the 3 Mb region (Figure 4) to suggest potential regulatory relationships between identified variants and nearby genes. The top candidate variant was located within the same TAD as *FOXG1*, and hereafter this TAD will be referred to as the FOXG1-TAD. Both *FOXG1* and the sequence surrounding the candidate variant are highly conserved across vertebrates, with the candidate sequence retained from fish to humans (Figure 6).

**Figure 4.**
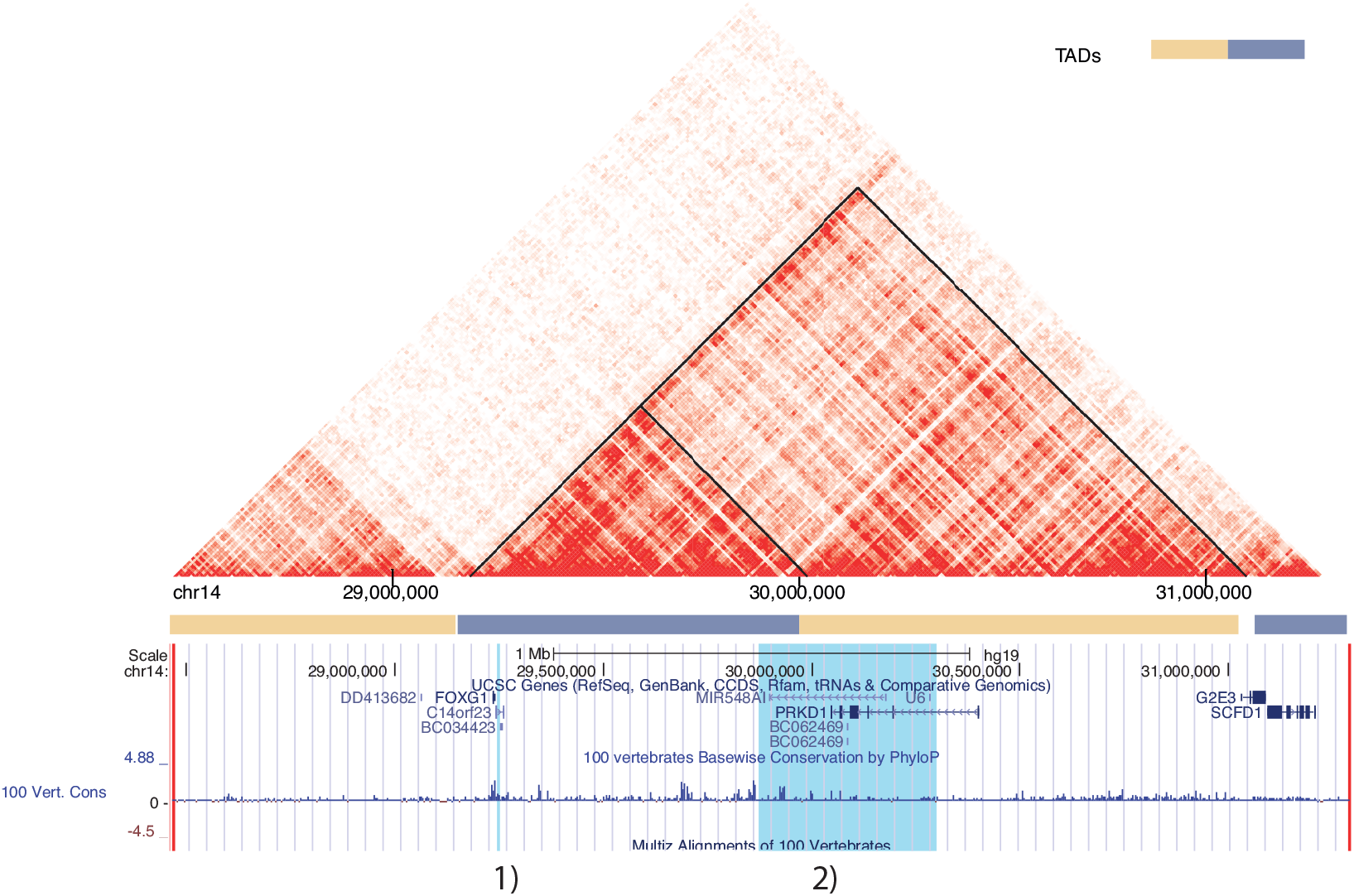
Topologically associating domains within the 3 Mb core region. This heatmap illustrates the chromatin interaction based on Hi-C data (41). The deeper the red color, the stronger the intra-chromosomal interaction between corresponding segments of the DNA. FOXG1-TADs are indicated by the black triangle shapes. Three blue highlights from left to right correspond to the putative regulatory region within the FOXG1-TAD: 1) chr14:29247628 TAAAC > T 2) The SRO (smallest region of deletion overlap) regulation region affecting FOXG1 expression (21)

**Figure 5.**
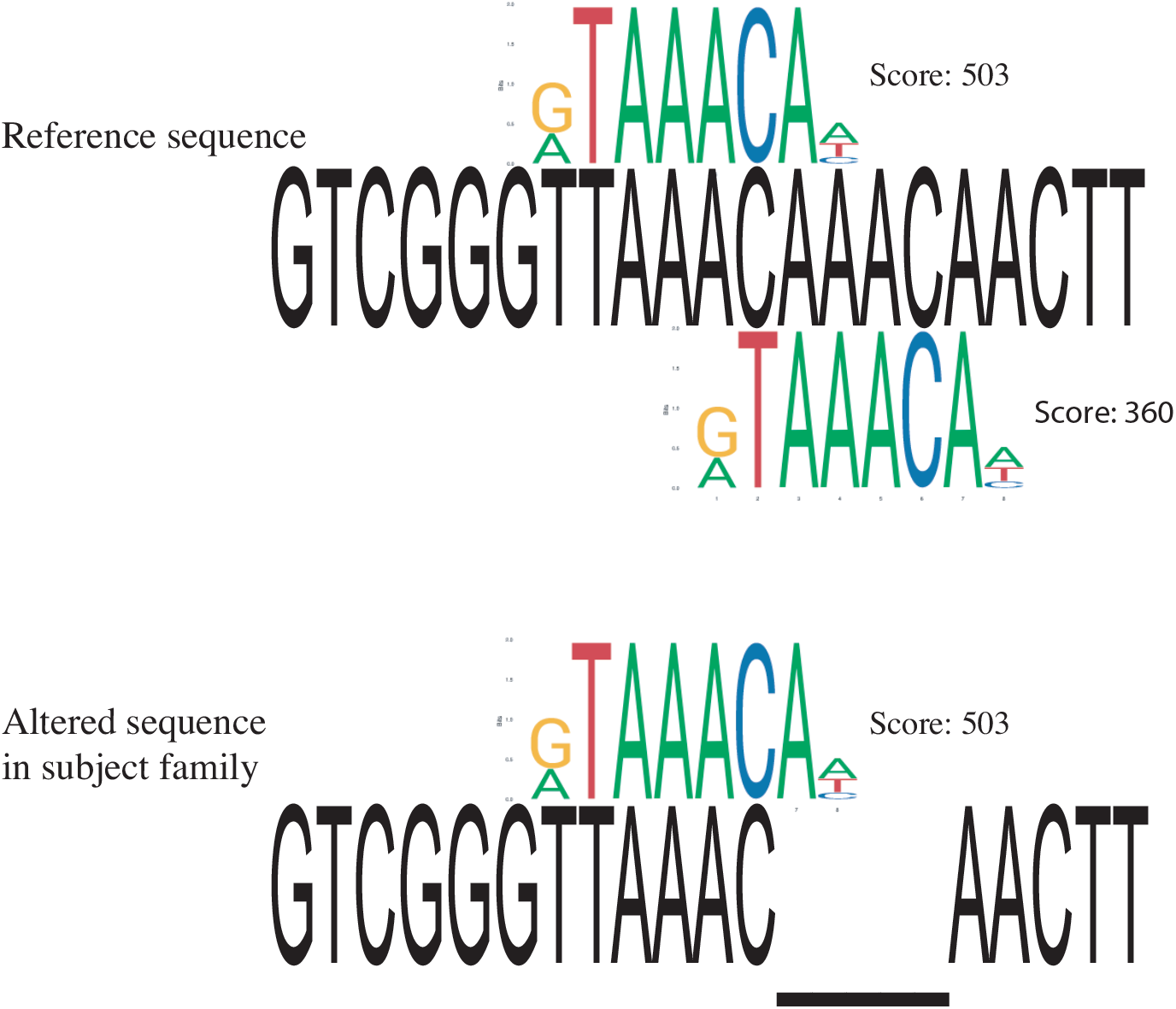
FOXG1 transcription factor binding site matching to reference and alternative sequence. Two FOXG1 TFBS are identified in reference sequence with scores of 503 and 360 respectively. Only one FOXG1 TFBS is identified in sequence with the 4 bp deletion. Scores are based on PWMScan with “JASPAR CORE 2018 vertebrates” library (Ambrosini G., PWMTools, http://ccg.vital-it.ch/pwmtools).

**Figure 6.**
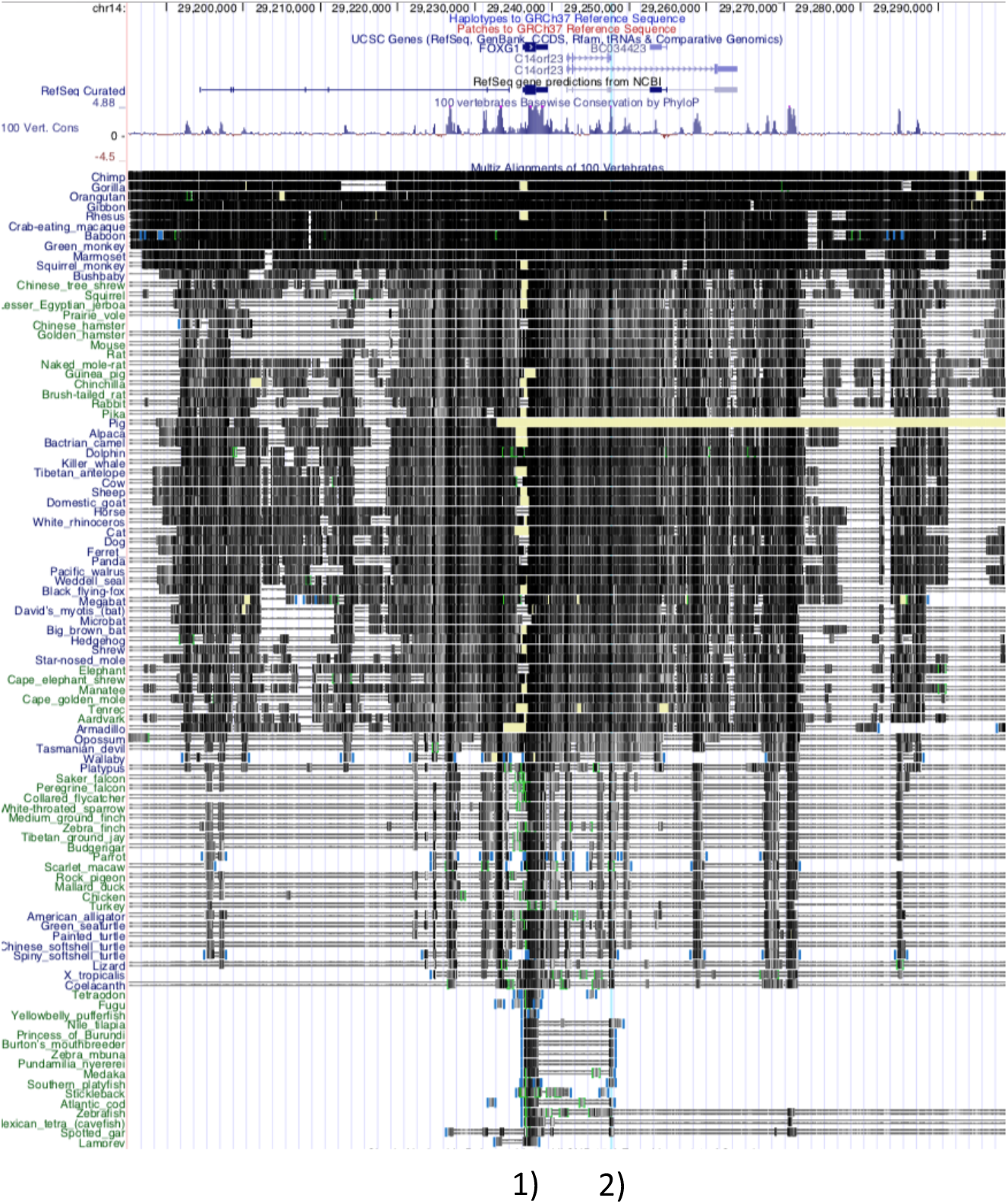
Ultra-conservation regions. 1) FOXG1 2) The 4bp deleted region

In human genome annotations, the variant chr14:29247628 TAAAC>T was located within an alternative exon of a long non-coding RNA gene (*LINC01551*). Within the mouse, chicken, and zebrafish annotation and supporting data, there were no RNA transcripts containing the variant (17). As the variant position is conserved back to fish, and the transcript evidence is not supportive of transcription of the highly conserved region in other species, we considered whether the variant might be situated within a *cis*-regulatory region. We examined predicted transcription factor binding motifs overlapping the deletion and observed a match to Forkhead transcription factor binding sites, including JASPAR profile matches for FOXC1, FOXI1, and FOXG1 (Figure 5). Proteins in the evolutionarily conserved superfamily of the Forkhead transcription factors share the presence of a DNA binding domain and a transactivation or transrepression effector region, and play a central role during development as well as in the adult (18). The binding motif for both FOXC1 and FOXI1 profiles would be obliterated by the deletion. Due to two consecutive AAAC repeats, the FOXG1 binding motif is present twice in the reference sequence, with one copy remaining after the deletion. Publicly available mouse ChIP-seq data (GSE96070) showed that Foxg1 binds to this site in cortex tissue from E14-15 brain. Thus, it appears that the deletion is situated within a Forkhead TF binding site, in a highly conserved region with conservation patterns consistent with a functional role in the *cis*-regulation of *FOXG1*.

## Discussion

We identified a new locus for isolated strabismus in a family, and this locus overlapped with the locus for FOXG1 syndrome, which has a high prevalence of strabismus. This finding suggests that the strabismus phenotype within FOXG1 syndrome can be isolated from the other phenotypes. In-depth phenotyping of individuals without strabismus surgery illustrated clinical heterogeneity of strabismus within the family, suggesting that while a specific molecular lesion may lead to strabismus, the specific clinical type is determined by other factors. We examined both coding and non-coding variants, which led to identification of a potential strabismus causing sequence alteration of a Forkhead TFBS within the FOXG1-TAD, suggesting disruption of *cis*-regulation.

To the best of our knowledge, our report contains the largest isolated strabismus pedigree in the literature with the highest LOD score. A LOD score of 4.69 was obtained for a single linkage peak on chromosome 14. Moreover, the segregation of the disease in this family is consistent with autosomal dominant inheritance. A 3 Mb haplotype was shared by all affected participants and was absent from unaffected participants.

In this family the strabismus types were not uniform. The various types of strabismus, however, appear to be caused by the same genetic factor since the linked haplotype is shared by all strabismic individuals. Our observation of strabismus variability in the subject family, review of the literature (19,20), and personal communication with other research groups suggests the current strabismus classification scheme is unhelpful for genetic studies: existing strabismus classification systems may inappropriately be separating individuals sharing a common underlying genetic cause and thus weaken study power. This may explain the paucity of studies detecting and confirming strabismus loci in the literature (3).

Since different types of strabismus appear to arise from the same genetic variant, a more reliable or nuanced classification system may be required. In phenotype ontologies, hierarchical classification systems allow for groupings to be examined at multiple levels of resolution. This important capacity is missing in our current approach to strabismus phenotyping. An expansion of the classification system could lead to an improved clinical understanding of strabismus and more efficient strategies to study the genetics of strabismus.

This 3 Mb region overlaps with microdeletions/microduplications known to cause FOXG1 syndrome in which a high prevalence of strabismus is observed. FOXG1 syndrome, which is also known as congenital Rett syndrome, is a neurological disorder characterized by impaired development and structural brain abnormalities. Strikingly, 84% of affected individuals display strabismus (5). Distal microdeletions that disrupt the topologically associating domains can lead to FOXG1 syndrome while *FOXG1* remains intact, indicating that mis-regulation of *FOXG1* can cause phenotypic change (21). Due to the close proximity and shared TAD with *FOXG1*, the 4bp deletion is speculated to alter *FOXG1* expression (Figure 4). A spectrum of partially overlapping phenotypes have been reported in patients with FOXG1 syndrome (5). The separability of the strabismus phenotype from intellectual disability and other severe disabling phenotypes therefore represents an important insight.

Close examination of diverse data provided important insights into the potential regulatory impact of the 4bp deletion. First, the sequences surrounding the deletion are highly conserved in the genomes of vertebrates, suggesting that it is under evolutionary selection and that a change may be more likely to have a functional impact. Indeed, the sequence containing the deletion and the coding region of *FOXG1* were the only two highly conserved elements in a 180 kb neighborhood (Figure 6). In addition, this conserved sequence was not supported as being part of a long non-coding RNA in other species (e.g. mouse, chicken, frog), implying a *cis*-regulatory effect. Second, the conserved sequence disrupted by the deletion was predicted to be a TFBS for Forkhead transcription factors, including FOXG1, according to the binding site profiles from JASPAR (22). Third, Foxg1 ChIP-seq data from E14-15 mouse brain (GSE96070) showed that Foxg1 bound to this sequence.

The binding of Foxg1 to this sequence in mouse provides the basis for the hypothesis of disrupted FOXG1 auto-regulation leading to strabismus in the subject family. The proposed auto-regulatory model is illustrated in Figure 7. *FOXG1* is transcribed and translated, the transcription factor binds to the target sequence, helping to maintain the appropriate expression of *FOXG1* during critical developmental period. The disruption of the FOXG1 binding site leads to dysregulation of *FOXG1* expression in a highly specific developmental context that results in the isolated strabismus phenotype.

**Figure 7.**
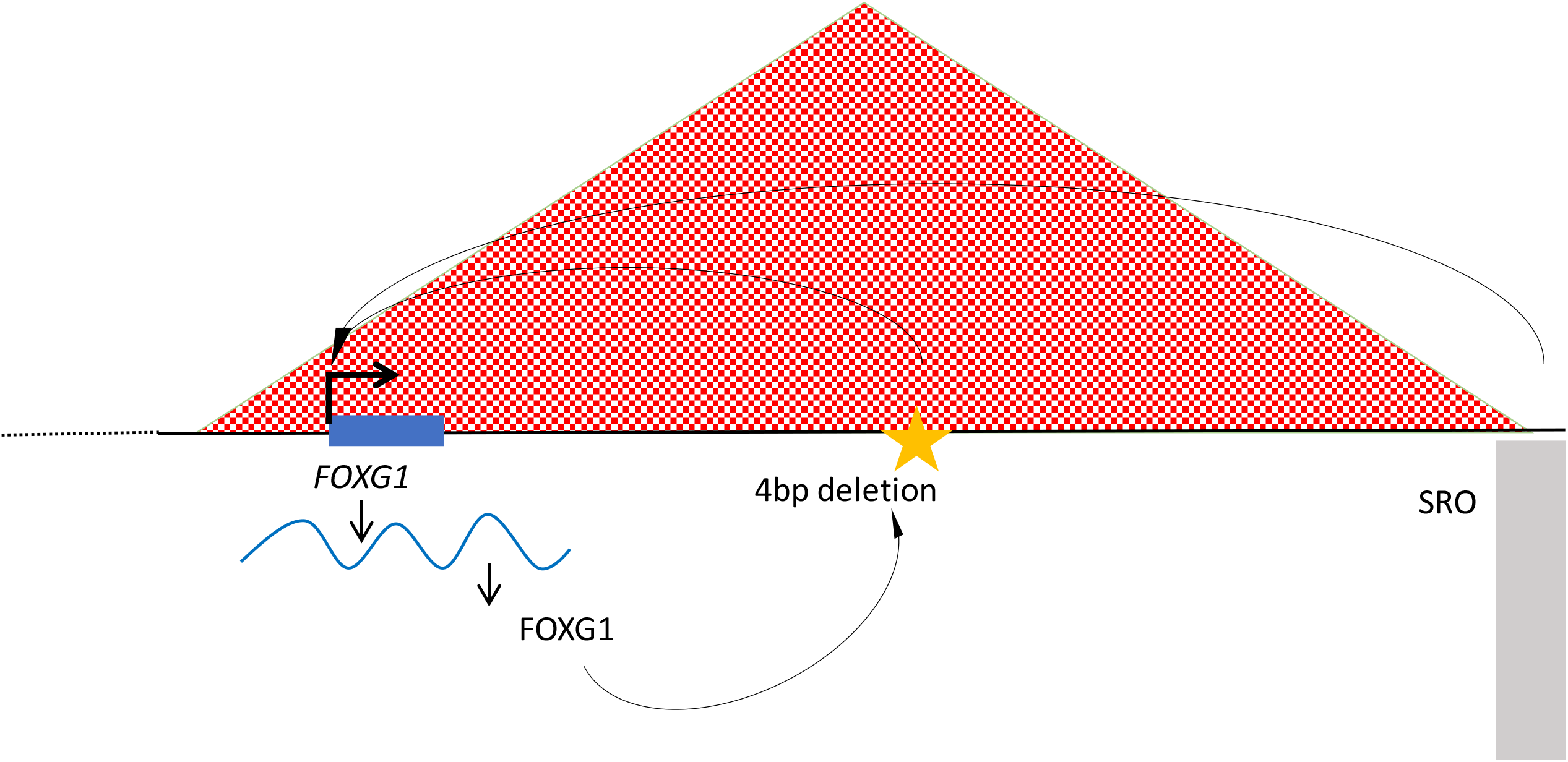
Cis-regulatory mechanism within FOXG1-TAD. 1) 4bp deletion chr14:29247628 TAAAC > T 2) The SRO (smallest region of deletion overlap) regulation region affecting FOXG1 expression (21)

Auto-regulation for key developmental transcription factors in vision is not new to the field. The SIMO regulatory sequence controlling expression of the *PAX6* transcription factor gene is an example of such a distal auto-regulatory element (23). While Pax6 is a crucial transcription factor for delineating the dorsal forebrain in mouse E10.0, Foxg1 is a critical transcription factor for delineating the ventral forebrain in mouse E9.0 (24). Thus, they may share similar sensitivity to regulatory disruption.

FOXG1 expression is strongest in fetal brain and its dysregulation leads to unbalanced development of excitatory and inhibitory synapses in iPSC-derived neurons and mice (25). In combination with other transcription factors, Foxg1 in pyramidal neurons is crucial for establishing cortical layers and axon trajectory of callosal projection neurons. Moreover, some Foxg1-directed processes are more vulnerable to dosage changes than others (26). These observations suggest that Foxg1 has a dosage and time sensitive role in different brain structures. This implies that an alteration in Foxg1 expression pattern can have a very specific impact, and the specific phenotype can be separable from the rest.

In summary, we identified a 3 Mb region on chromosome 14 that is linked to autosomal dominant transmission of isolated strabismus. The region contains *FOXG1*, which has been previously associated with strabismus in 84% of patients with syndromic disruptions. Within the 3 Mb region, the top candidate variant is situated within a FOXG1 transcription factor binding motif, suggesting that disrupted auto-regulation could be the mechanism underlying the observed strabismus phenotype. As the causal functional alteration remains to be proven, additional studies will be required to identify other families with genetic forms of strabismus mapping to the locus and to conclusively prove the causal sequence alteration and its pathophysiological mechanism.

## Materials and Methods

### Patient ascertainment

The study was approved by the University of British Columbia Children’s & Women’s Research Ethics Board (approval number CW10-0317/H10-03215), and written consent forms were obtained from all 14 participants. A seven-generation pedigree was constructed based on family records, including photos displaying eye alignment. Thirteen participants were descendants of a common ancestor; nine of them reported early onset isolated strabismus, and the other four reported no strabismus. The common ancestor was reported to be of European origin.

Except for 006 and 005, the other eight affected descendants and four unaffected descendants were examined by one or more of three ophthalmologists (Drs. J. Horton, V. Pegado, and C. Lyons). All participants were asked about the age of onset (if applicable), ocular history, and medical history. Examination included visual acuity, pupil observation, eye movements, ocular alignment, stereopsis, slit lamp examination, fundus examination, and intraocular pressure. Individuals 009, 011, and 013 did not have a history of extraocular muscle surgery and therefore underwent full orthoptic exams. Subjects were asked about other medical or physical characteristics, with no reports spanning beyond immediate nuclear family members.

### DNA isolation

Genomic DNA of participants was isolated from either saliva or blood. At least 4 ml blood samples or 6 ml saliva samples were collected for one round of next generation sequencing, and at least a 2 ml saliva sample was collected from participants for array genotyping. Blood samples were collected in a clinical setting while saliva samples were collected using Oragene-DNA (OG-500) saliva kits. DNA was extracted from blood samples using the Qiagen QIAsymphony SP instrument and the QIAsymphony DNA Midi Kit and from saliva samples with DNA Genotek prepIT-L2P sample preparation kit following protocol # PD-PR-015. Approximately, 7-10 µg DNA per sample at a concentration no less than 70 ng/µl was sent for sequencing. A 500 ng DNA per sample at a concentration of at least 50 ng/µl was sent for array genotyping.

### Genotyping: statistical linkage analysis and haplotype analysis

Genotyping was performed by The Centre for Applied Genomics, The Hospital for Sick Children, Toronto, Canada. The assay was performed on HumanOmni2.5-8v1_C, using the Infinium LCG assay (Illumina). Standard quality control steps were performed on the genotypes, including sex check, call rate, autosomal heterozygosity, and verification of the pedigree structure. Simulations were performed to determine the maximum possible LOD (logarithm of the odds) score for different model parameters under the alternative hypothesis (linkage). SLINK (27) was used to simulate pedigrees under dominant and recessive models with a range of disease allele frequencies and penetrance. For a particular model, the maximum LOD score from the analysis of 1000 simulated pedigrees was declared the maximum LOD score.

Multiple filters were applied to select a set of markers suitable for linkage analysis. Only markers with alleles unambiguous for strand information on the autosomes and X chromosomes were kept. Genotype data from HapMap3 European populations (28) were used to estimate marker allele frequencies, and a minor allele frequency >0.45 and pairwise r^2^ < 0.1 were selected. A set of 17,779 SNPs was obtained after a SNP filtering step. Merlin 1.1.2 (29) was used to perform multipoint linkage analysis under the same model as in the SLINK simulation. Analysis of the X chromosome was performed within Merlin using the standard procedures (29).

As individual 014 was recruited at a later time point, we extracted single nucleotide polymorphisms (SNPs) from whole genome sequencing (WGS) data. We then used the combined SNP genotypes for linkage analysis on the family and performed genome-wide parametric and non-parametric linkage analyses using Merlin 1.1.2. To refine the boundaries of the linked region, we examined the SNPs from both edges (rs714641 and rs1951187) manually between descendants and identified the minimum shared haplotype region.

### Whole-exome sequencing

We performed whole exome sequencing (WES) on 001 and 011, affected third cousins. WES was performed via the Agilent SureSelect Human All Exon 38Mb kit and Illumina HiSEQ 2000 platform (performed by Perkin Elmer) with an average coverage of 27X. The genomic aligners, Bowtie (version 0.12.9) and BWA (version 0.6.1), were used to map the paired-end reads to the hg19 reference genome (30,31). The Genome Analysis Toolkit (GATK) (version 1.0) performed local re-alignment, which allowed for correcting misalignment at the extremity of reads (32).

SAMtools (versions 0.1.18) was applied to call variants from aligned WES reads (33). In-house scripts were used to filter variants according to the following criteria: under an autosomal dominant model, with a frequency not higher than 1% in dbSNP build 135, non-synonymous coding variants, and predicted by SIFT (12) to be ‘damaging’ or indeterminate.

### Whole-genome sequencing

Two rounds of WGS were conducted as the project progressed. WGS was first performed on 001, 013, and 014, who were three distantly related affected cousins, on an Illumina HiSEQ 2000 platform (BGI America) generating paired-end reads of 125 bp and average coverage of 37X. An informatics pipeline (similar to the WES pipeline, but with newer versions of software) was applied to this set of WGS data: Bowtie (version 1.0.0) and BWA (version 0.7.5a) for mapping the paired-end reads to the hg19 reference genome (30,31), GATK (version 2.8) for local re-alignment (32), and SAMtools (version 0.1.19) for variant calling (33).

Variants located within the linkage region were selected for further analysis. Allele frequency was assessed using dbSNP build 137 and Exome Variant Server (URL: http://evs.gs.washington.edu/EVS/) and variants with a frequency higher than 1% were excluded. Heterozygous variants shared across the three samples were selected, and SnpEff (34) (with hg19 database) was applied to annotate those variants.

Later, in order to obtain PCR-free results suitable for analysis of short tandem repeats, WGS was performed on 011, 012, and 013, a trio, on an Illumina NovaSeq platform (Macrogen) with an average coverage of 45X. A different informatic pipeline was applied to this set of WGS data: BWA mem (version 0.7.12) for mapping the paired-end reads to the GRCh37 reference (http://www.bcgsc.ca/downloads/genomes/9606/hg19/1000genomes/bwa_ind/genome/), SAMtools (version 1.2) for file format conversion, duplicate marking with Picard (version 1.139), GATK for local re-alignment (version 3.4-46), and GATK HaplotypeCaller for variant calling (version 3.4-46). Variants were soft-filtered using BCFTools (version 1.8) keeping variants with at least 10 reads supporting the alternate allele and a max depth of 300. Filtered variants were then annotated and normalized using SnpEff (version 4.11; gene version GRCh37.75), VT (version 0.5772), and VCFAnno (version 0.2.8) (35). Filtered, annotated variants are then converted into a GEMINI database (version 0.19.1) (36) using VCF2DB (https://github.com/quinlan-lab/vcf2db). Specific GEMINI queries were performed for variants under the autosomal dominant model, with details below. Scripts for processing the data, and details regarding databases annotated against using VCFAnno and CNV analyses can be found online (https://github.com/Phillip-a-richmond/AnnotateVariants/tree/master/Strabismus) (37). Variants under the autosomal dominant model (shared between 011 and 013) located within the linkage region were selected for further investigation. Reflecting the upper end of reported strabismus population frequency, all variants with allele frequency <4% were manually reviewed. However, it is expected that dominant transmission of isolated strabismus is a rare event, and therefore the results reported in this manuscript pertain to those variants with allele frequency <1% and <10 homozygous individuals for the minor allele in gnomAD 2.0 (10). Candidate rare variants within the region were then assigned to gene and then manually examined for potential molecular impact of the gene by database searches of tissue expression patterns and previously published data.

### Non-coding variant annotation and interpretation

To enable analysis of non-coding variants, we used multiple databases and corresponding bioinformatic tools to annotate such variants, including functional annotation of the mammalian genome 5 (FANTOM5) database (38), JASPAR (22), Segway (39), RegulomeDB (40) and Combined Annotation Dependent Depletion (CADD) (14). Data related to the candidate variants were visualized in the UCSC Genome Browser. The 100 Vertebrates Basewise Conservation PhyloP track was used for examining the conservation status. Due to the incomplete annotation of noncoding RNAs across different species, Emsembl BLAST/BLAT was used to find potential orthologs in other vertebrates, and RefSeq genes and ESTs were considered as evidence for RNA transcripts (17). Based on the qualitative assessment, the top prioritized variant was confirmed to be present by Sanger sequencing in all of the affected subjects and absent from unaffected subjects.

## Data Availability

The genome wide sequencing data cannot be shared publicly because of privacy concern.

## Acknowledgement

We thank Dr. Jonathan C. Horton (University of California San Francisco) for generously offering his time to examine multiple participants in his clinic. We thank Ms. Dora Pak for research management support. Funding for X.C.Y. is provided by a Frederick Banting and Charles Best Canada Graduate Scholarship (GSD-146285) from the Canadian Institutes of Health Research, BCCHRI-CIHR-UBC MD/PhD Studentship, and NSERC Discovery Grant (RGPIN-355532-10, RGPIN-2017-06824). The SNP genotyping was performed under the Care4Rare Canada Consortium funded by Genome Canada and the Ontario Genomics Institute (OGI-147), the Canadian Institutes of Health Research, Ontario Research Fund, Genome Alberta, Genome British Columbia, Genome Quebec, and Children’s Hospital of Eastern Ontario Foundation. C.J.R is supported by a Michael Smith Foundation for Health Research Scholar Award.

## Supporting Information

**Supplementary Figure 1.**
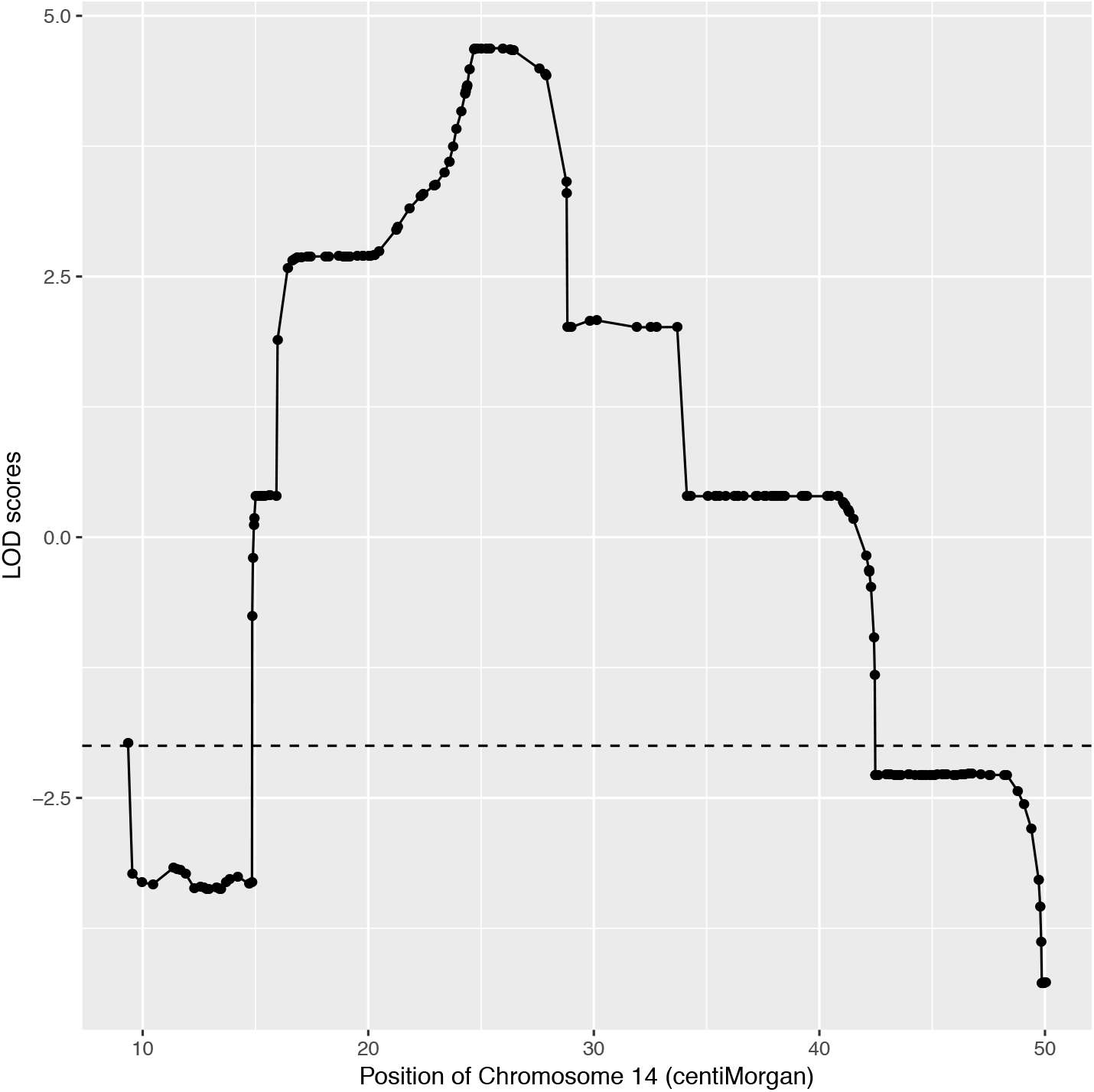
LOD score plot for the observed linked region on chromosome 14. The horizontal axis represents the location in terms of centiMorgans. The displayed region spans the physical positions of chr14:28,467,136 – 30,045,978 (hg19/GRCh37). The vertical axis represents LOD score. The dash line indicates the threshold of LOD <-2.

**Supplementary Figure 2.**
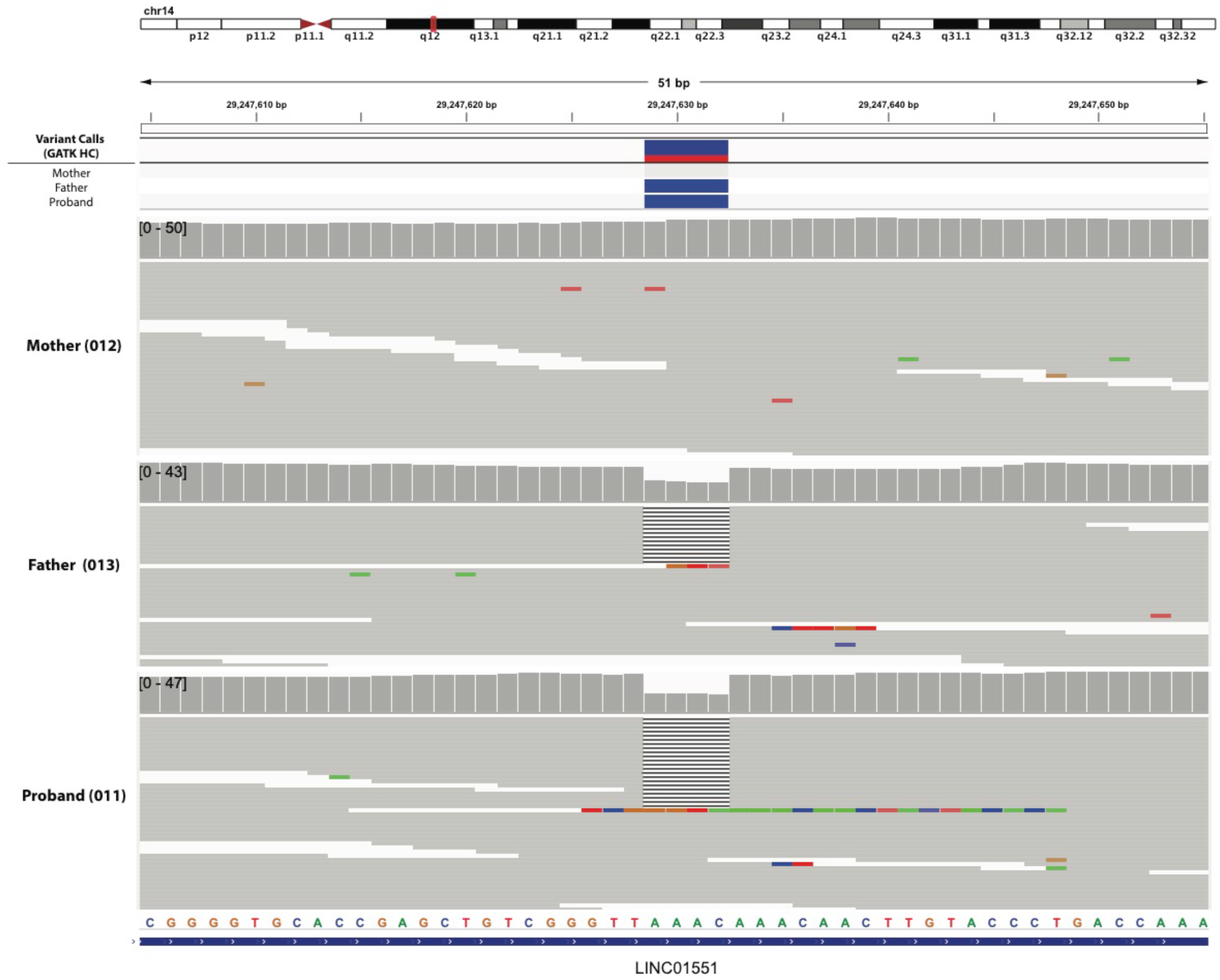
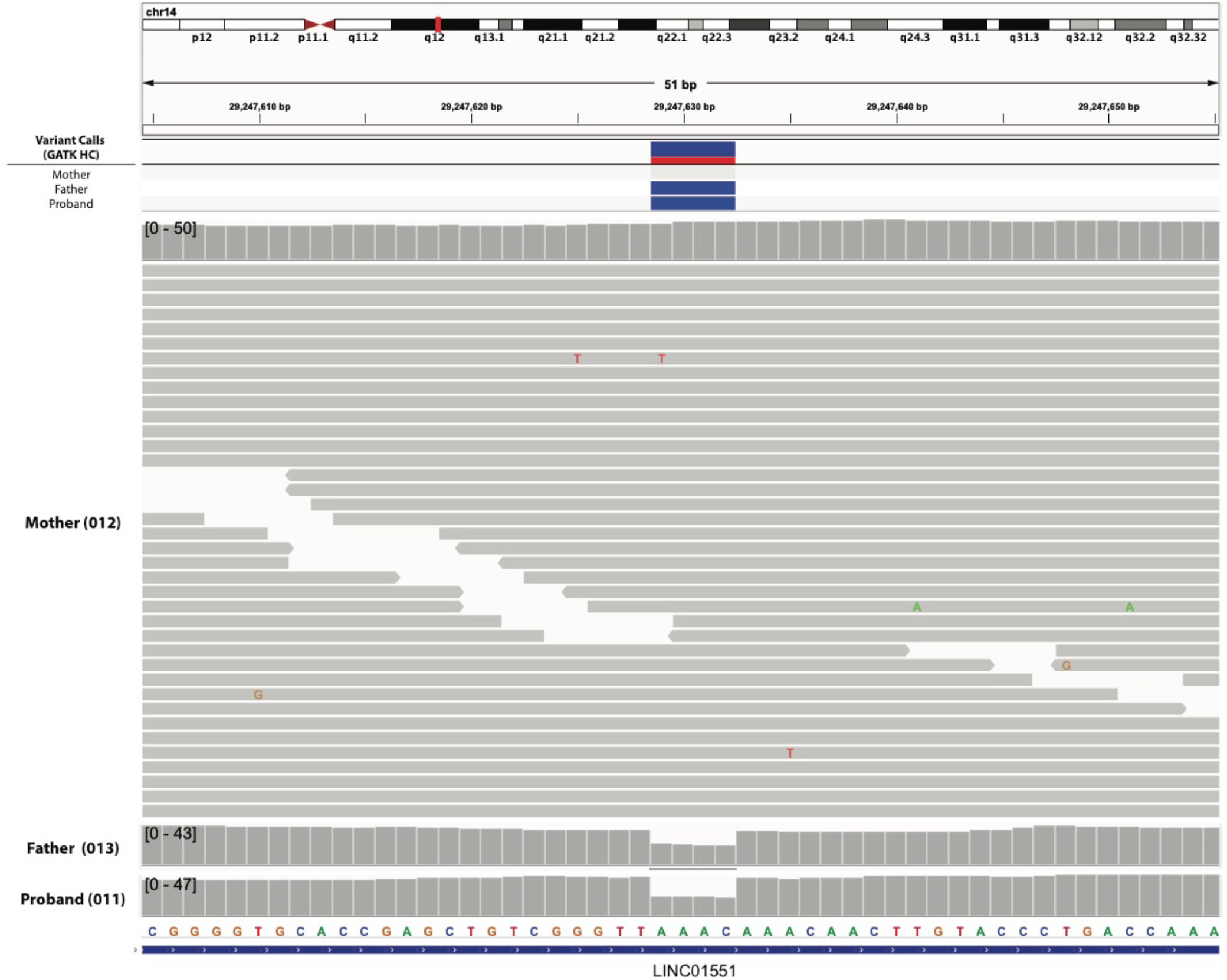
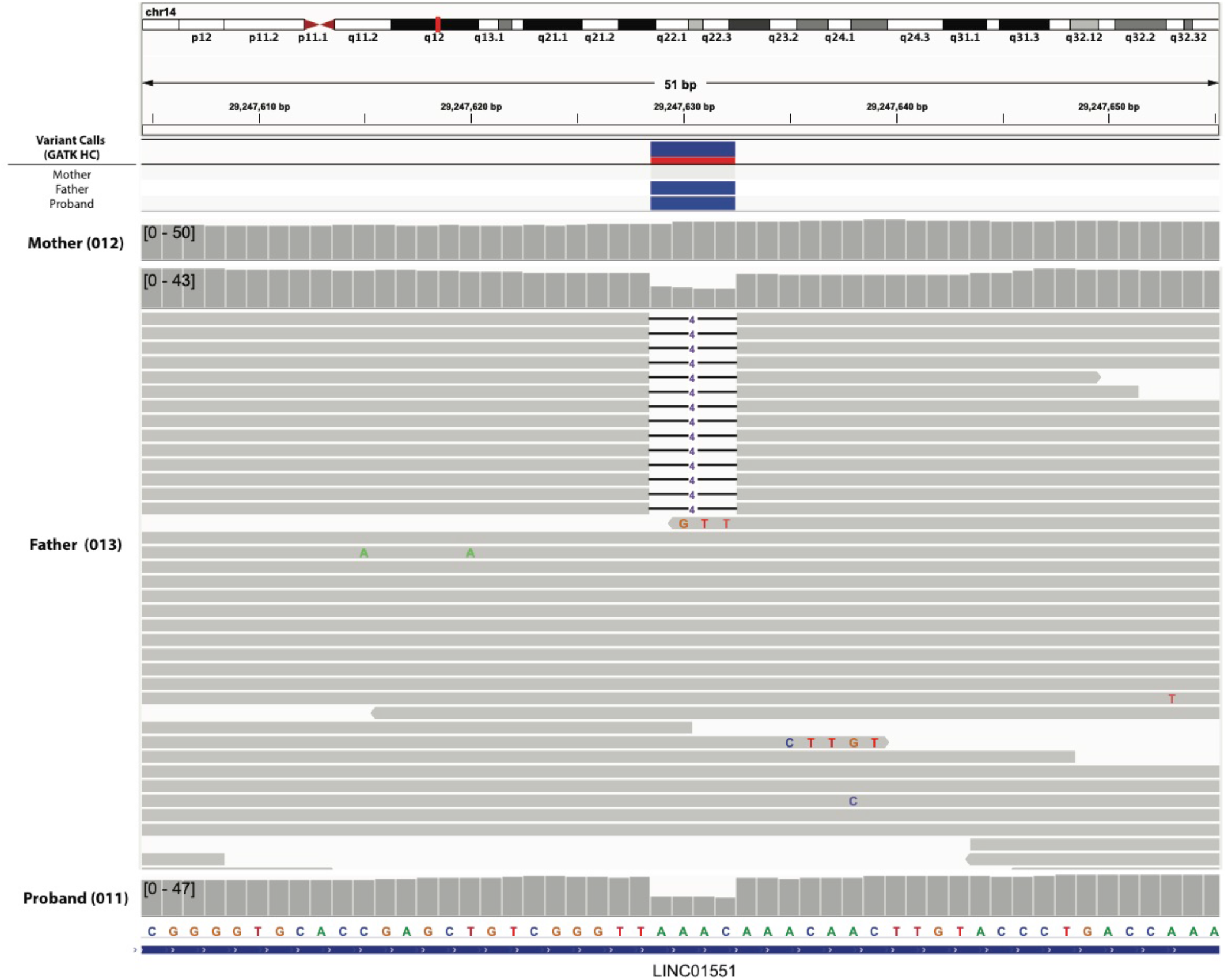
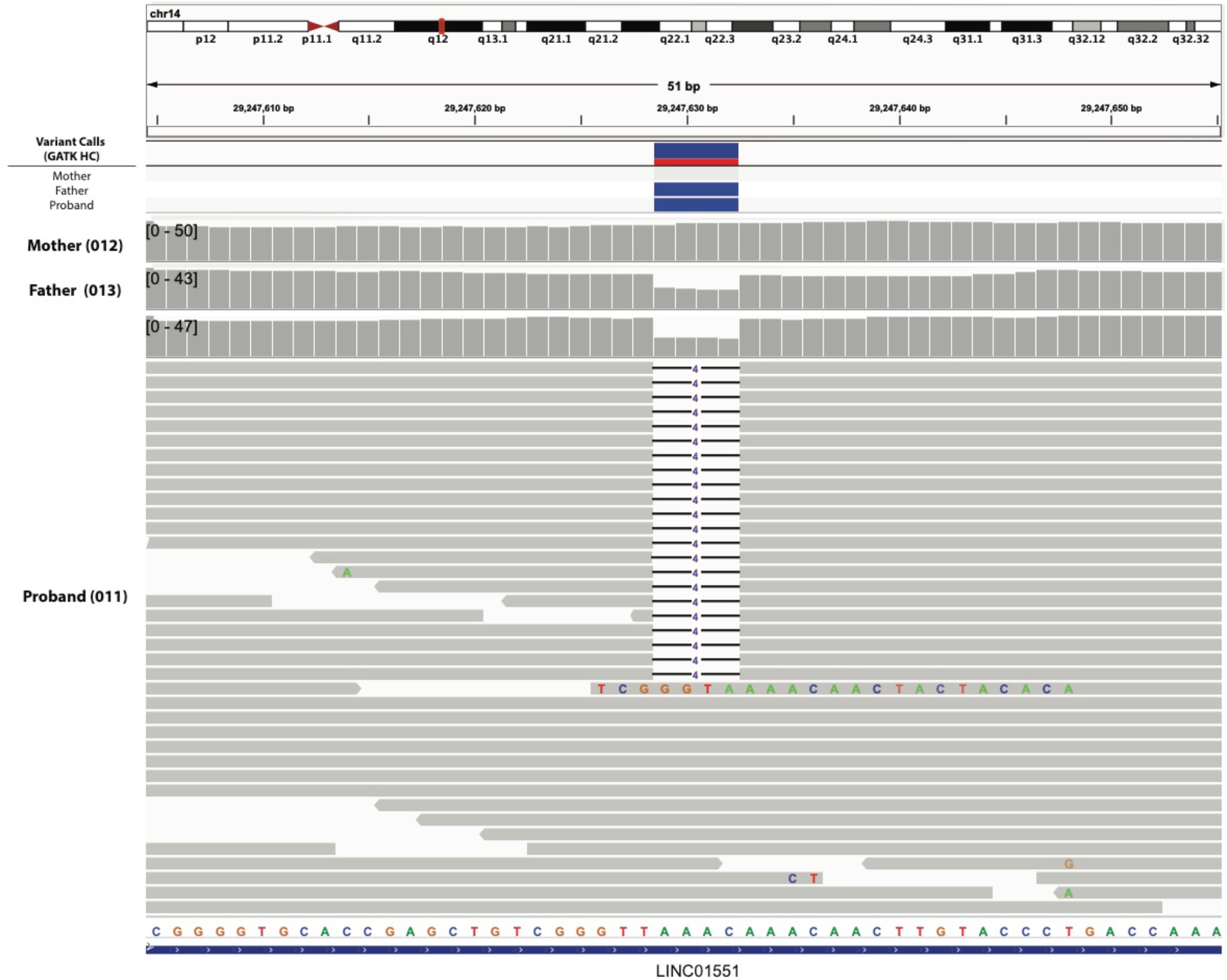
a) Integrative Genomic Viewer (IGV) screenshot of the WGS for the parent-child trio. (Affected: 011 and 013; Unaffected: 012). The horizontal axis represents the sequence of the reference genome. A small portion of the aligned reads for each of the three individuals is shown as indicated by the labels along the vertical axis. b) Extended view of IGV screenshot for 012 (Mother). c) Extended view of IGV screenshot for 013 (Father). d) Extended view of IGV screenshot for 011 (Proband). (Detailed information about the IGV display can be obtained on https://software.broadinstitute.org/software/igv/)

